# Theoretical investigation of pre-symptomatic SARS-CoV-2 person-to-person transmission in households

**DOI:** 10.1101/2020.05.12.20099085

**Authors:** Yehuda Arav, Ziv Klausner, Eyal Fattal

**Affiliations:** Department of Applied Mathematics, Israeli Institute for Biological Research, PO Box 19, Ness-Ziona, 7410001 Israel

## Abstract

Since its emergence, the phenomenon of SARS-CoV-2 transmission by seemingly healthy individuals has become a major challenge in the effort to achieve control of the pandemic. Identifying the modes of transmission that drive this phenomenon is a perquisite in devising effective control measures, but to date it is still under debate. To address this problem, we have formulated a detailed mathematical model of discrete human actions (such as coughs, sneezes, and touching) and the decay of the virus in the environment. To take into account both discrete and continuous events we have extended the common modelling approach and employed a hybrid stochastic mathematical framework. This allows us to calculate higher order statistics which are crucial for the reconstruction of the observed distributions. We focused on transmission within a household, the venue with the highest risk of infection and validated the model results against the observed secondary attack rate and the serial interval distribution. Detailed analysis of the model results identified the dominant driver of pre-symptomatic transmission as the contact route via hand-face transfer and showed that wearing masks and avoiding physical contact are an effective prevention strategy. These results provide a sound scientific basis to the present recommendations of the WHO and the CDC.

## Introduction

The phenomenon of SARS-CoV-2 transmission by pre-symptomatic, otherwise seemingly healthy, individuals poses a major challenge for policy makers’ efforts to achieve control of the COVID-19 pandemic, as traditional health strategies rely on case detection through manifestation of symptoms^1^. However, the mechanism that enables this transmission is not fully understood. Generally, respiratory viruses such as SARS-CoV-2 propagate via four modes of transmission^2^: direct physical contact between people, indirect physical contact via intermediate object, droplets and droplet nuclei. Transmission by droplets and droplet nuclei is mediated by virus containing particles that were emitted when a person coughs, sneezes or speaks. The droplets travel less than 1.5m ^3^, due to their size, and settle on the facial membranes of nearby individuals or on surfaces. Droplet nuclei remain suspended in the air and may infect a susceptible individual once they penetrate the respiratory tract. The commonly accepted cutoff between droplets and droplet nuclei is 5*µm* ^2^. However, Xie et al.^3^ showed that droplets that are smaller than approximately 100*µm* evaporate to their nuclei size before reaching the ground.

The relative contribution of the different modes of transmission in indoor environments is still under debate^4–9^. The controversy revolves about the relative importance of the droplet nuclei mode of transmission. Several studies have argued that the transmission of the SARS-CoV-2 virus is mediated primarily by close and unprotected contact (e.g., via physical contact and droplets)^4–7^, while others have argued that breathing droplet nuclei is the main mode of transmission^8, 9^. The close contact transmission hypothesis relies on the analysis of COVID-19 cases^6^ and the relatively low secondary attack rate (SAR, the probability of an infected person to infect a susceptible person) of 10%-16% that was observed in households^5, 10–13^. The droplet nuclei hypothesis relies on several theoretical investigations^8, 9^. The attempts to identify SARS-CoV-2 in air sampling taken from infection isolation rooms in hospitals and households yield conflicting results. Several studies^14–16^ found positive samples while others^17–19^ reported negative air samples (for example, Dohla et al.,^19^ reported negative air samples that were taken from households).

The aim of this study is to quantify the relative contribution of the different modes of transmission of SARS-CoV-2 to infection by pre-symptomatic individuals. We focus in this study on the transmission within a household environment, the venue with the highest risk of infection^5, 10^. The approach taken here is an integrative detailed mechanistic modelling that describes explicitly the transfer of SARS-CoV-2 between individuals in different modes of transmission, similar to the approach used by Nicas and Sun^20^ and by Atkinson and Wein^21^ for quantifying the modes of transmission of respiratory viruses. In this work we have extended the mathematical framework of Atkinson and Wein^21^ to take into account random discrete human actions (such as coughs, sneezes and contact with objects and other people), rather than considering only the mean kinetics. This was achieved by employing a hybrid stochastic mathematical framework which allows us to explicitly calculate higher order statistics which are crucial for the reconstruction of the observed distributions. Following this, the model is validated by reconstruction of observed fundamental attributes of the pandemic, the secondary attack rate (SAR) and the serial interval distribution. Then, the model is used to assess the contribution of each of the transmission modes as well the effectiveness of different prevention measures.

### Outline of the Mathematical model

The model presented in this study incorporates the processes that influence the number of SARS-CoV-2 virus particles transferred from a pre-symptomatic infected individual, henceforth the primary, to a susceptible individual, henceforth the secondary, and the probability to become infected (Figure 1).

**Figure 1.**
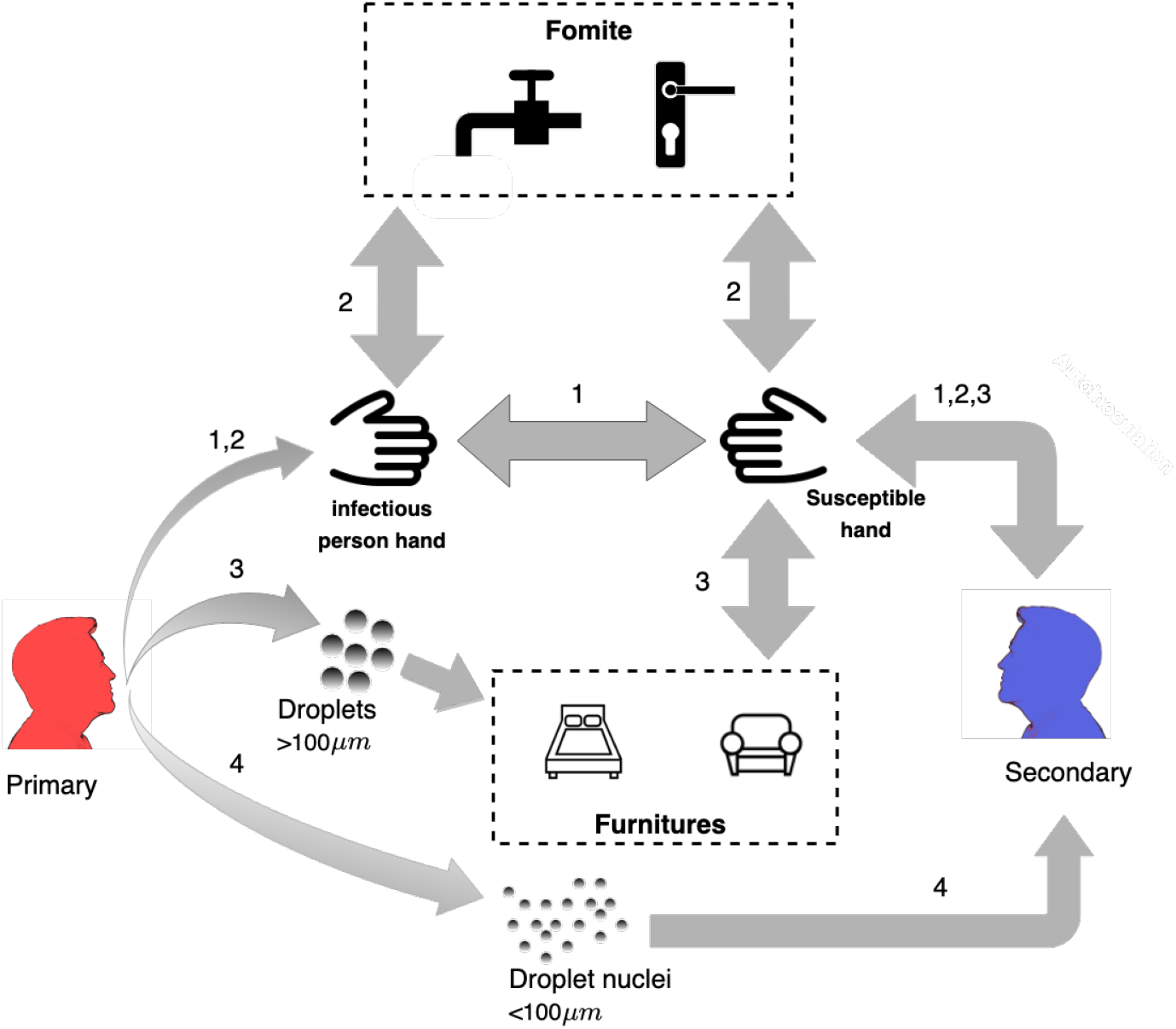
Schematic representation of the modes of transmission from the primary (infector) and secondary (infectee) individuals. (1) Direct contact (2) Indirect contact via fomites (3) Indirect contact via surfaces (4) droplet nuclei.

Contact transmission begins when the primary touches his facial membranes and, as a result, contaminates his own hands. Then, the primary transfers the virus either through direct physical contact (Figure 1, mode 1) or indirectly via small frequently touched object (fomits), such as a doorknob or a faucet, (Figure 1, mode 2) to the hands of the secondary. Eventually, the secondary places his hands into nose, mouth or eyes, which might cause an infection^21, 22^. The droplet and droplet nuclei modes of transmission (Figure 1, modes 3 and 4, respectively), begin when the primary coughs, sneezes, or speaks and expels virus containing droplets. Droplets larger than 100*µm* settle by gravity within 1.5*m* ^3^ and contaminate large surfaces such as furniture and table tops (environmental surfaces), while smaller droplets dry out and form droplet nuclei which remain suspended in the air. As a result, the droplet nuclei may be carried over distances greater than 1.5 m by the air currents of the room^3^. The deposition of droplets directly on the mocusa of close contacts is a rare event in workplace or household settings^21^. Therefore, we have considered here only the contamination of environmental surfaces by the droplets after they have settled. The contaminated areas on the environmental surfaces might also contaminate the hands of the secondary individual when he touches them. The probability of infection increases with the number of SARS-CoV-2 particles that reach facial membranes of the secondary individual.

The processes described in Figure 1 consist of both discrete random events which are the actions of the individuals, such as touching each other or touching the facial membranes and continuous events which consist of environmental processes. Hence, we used a hybrid continuous and stochastic-jump framework^23^ to describe the dynamics of the transmission and infection processes using a coupled system of differential equations. The actions of the individuals are described as stochastic jump Poisson processes, while the environmental processes are described using continuum dynamics. The model explicitly tracks the dynamics of the concentration on the hands of the individuals (equation 8), the concentration on the fomites (equation 9), concentration on environmental surfaces (equation 11) and the concentration of the droplet nuclei in the air (equation 10). A complete list of the model equations and values of the corresponding parameters are provided in the Methods section. Since the actions of the individuals are represented as a stochastic process, we have conducted a Monte Carlo simulation in which multiple realizations were computed to obtain the appropriate ensemble statistics. Using a Monte-Carlo simulation the probability distributions are embedded in the input parameters directly and allow the comparison of the model results to observed distributions. Each realization begins when the primary become infected and begins an incubation period whose duration is drawn from a log-normal distribution with a mean of 5 days and standard deviation (SD) of 0.45 days^24^. The viral load of the primary increases exponentially with time^21^ reaching a maximal level at the end of the incubation period^25^. During that time, the primary and secondary individuals perform a series of randomized actions such as touching fomites, touching environmental surfaces, coughing, sneezing, talking, touching each other, or each touching his own face. The probability that the secondary individual will be infected is determined from his accumulated exposure over a time interval (equation 12) using the dose-response curve that was reported for SARS-CoV-1^26^ (equation 7) and assumed to be similar to SARS-CoV-2. Each realization ends when the primary develops symptoms, in accordance with the public health policy that isolates the primary at the onset of symptoms.

We define a reference simulation as a simulation which corresponds to a normal, pre-symptomatic behaviour (parameters in Table 1).

**Table 1.** The hygienic and behavioral parameters of the reference simulation

| Parameters | Parameter Description | Value | Unit | Reference |
| --- | --- | --- | --- | --- |
| $\tau_{social}$ | Person to person physical contact frequency | 3 | 1/d | 27 |
| $\tau_{hand-face}$ | Rate of face touching | 0.2 | 1/min | 22 |
| $\tau_{hand-fomite}$ | Rate of fomite touching | 60 | 1/d | 28 |
| $\tau_{hand-furniture}$ | Rate of furniture touching | 1 | 1/min | 22 |
| $\tau_{hand-washing}$ | Rate of hand cleaning | 3 | 1/d | 28 |
| $\tau_{fomite-cleaning}$ | Rate of fomite cleaning | 2 | 1/d | 28 |

## Results

### Validation of the model

A necessary validation criteria for a model such as the one described in this study is to correctly simulate the distribution of the serial interval and the SAR. The serial interval is the time period between the symptoms’ onset in the primary and the secondary. Its distribution is closely associated with the estimation of the reproductive number and key transmission variables in epidemic models and is important for optimization of the length of the obligatory quarantine period and contact tracing strategies^29, 30^. The serial interval distribution of COVID-19 was estimated in many countries and was usually found to be gamma distributed with mean between 4.03 to 6.3 days and standard deviation between 3 and 4.2 days (Figure 2A, shaded area)^10, 31–33^.

**Figure 2.**
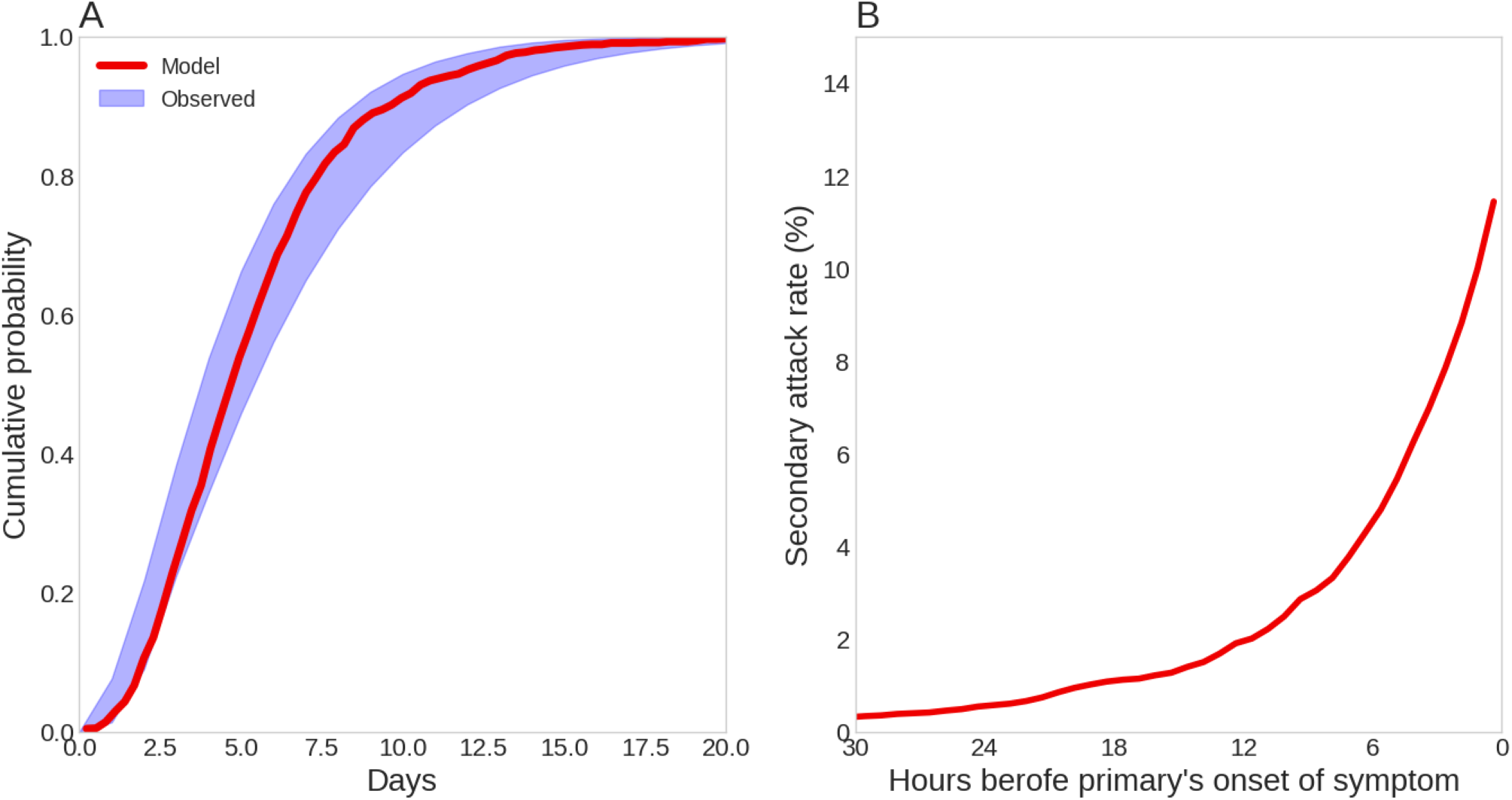
Model Prediction for the (A) Distribution of the serial index. Shaded area is the bounds of observed data^10, 31–33^ (B) The cumulative SAR over time.

The model prediction for the distribution of the serial interval and the SAR is obtained by conducting a Monte-Carlo simulation by solving equations 8 to 12 (see the Methods section) for 10000 realizations of the reference simulation (needed for convergence). Figure 2A compares the model predicted serial interval distribution (red line) with the distributions reported in the literature. As seen, the model prediction was well between the bounds of the different estimates of this distribution (Figure 2A).

As an additional validation, we compared the model prediction of the SAR to the values reported in the literature. The model predicts a SAR of 11.5% in the reference simulation, which is within the reported values ranging between 10 − 16%^5, 10–13^. We have also analyzed the contagious period of pre-symptomatic patients by examining the cumulative SAR over time (Figure 2B). As seen, the contagious period begins approximately 30 hours before the symptoms’ onset, with increasing probability of infection as the onset of the symptoms approaches. This result is consistent with the estimation of He, et al.^25^, that inferred from data of 77 transmission pairs (i.e., primary and secondary) a contagious period of approximately 2 days before symptoms’ onset.

Some of the parameters’ values were obtained from studies that also reported the range of variability of these values. Therefore, we have performed an extensive sensitivity analysis to check the robustness of the results (see Supplementary Information). The model’s results remain within the range of the values reported in the literature for the examined range of parameters.

### Modes of transmission in pre-syptomatic cases

Analyzing the realizations of the reference simulation, we have quantified the contribution of the different modes of transmission to the overall exposure in scenarios where the secondary was infected (Figure 3). Out of the total viral dose that was transmitted to the secondary, 64.5% (Inter quartile range, IQR: 55% − 80%) was received during direct contact events (mode 1) and 26% (IQR 13% − 32%) was received during indirect contact via fomites events (mode 2). The contribution of the large droplet route (mode 3) was negligible while the droplet nuclei transmission (mode 4), contributed 9.5% (IQR 3.6% − 12%) of of the total viral dose. Hence, according to our results, the contact mode of transmission (either direct or indirect) is the dominant mode of infection, accounting, overall, to the transfer of 90% of the viral dosage from the primary to the secondary. The main process that underlies the contact mode of transmission is the hand-face transfer. Therefore, hygienic and behavioral measures that operate on the elements that constitute the contact processes are expected to significantly reduce the risk of infection. These will be analyzed in the following section.

**Figure 3.**
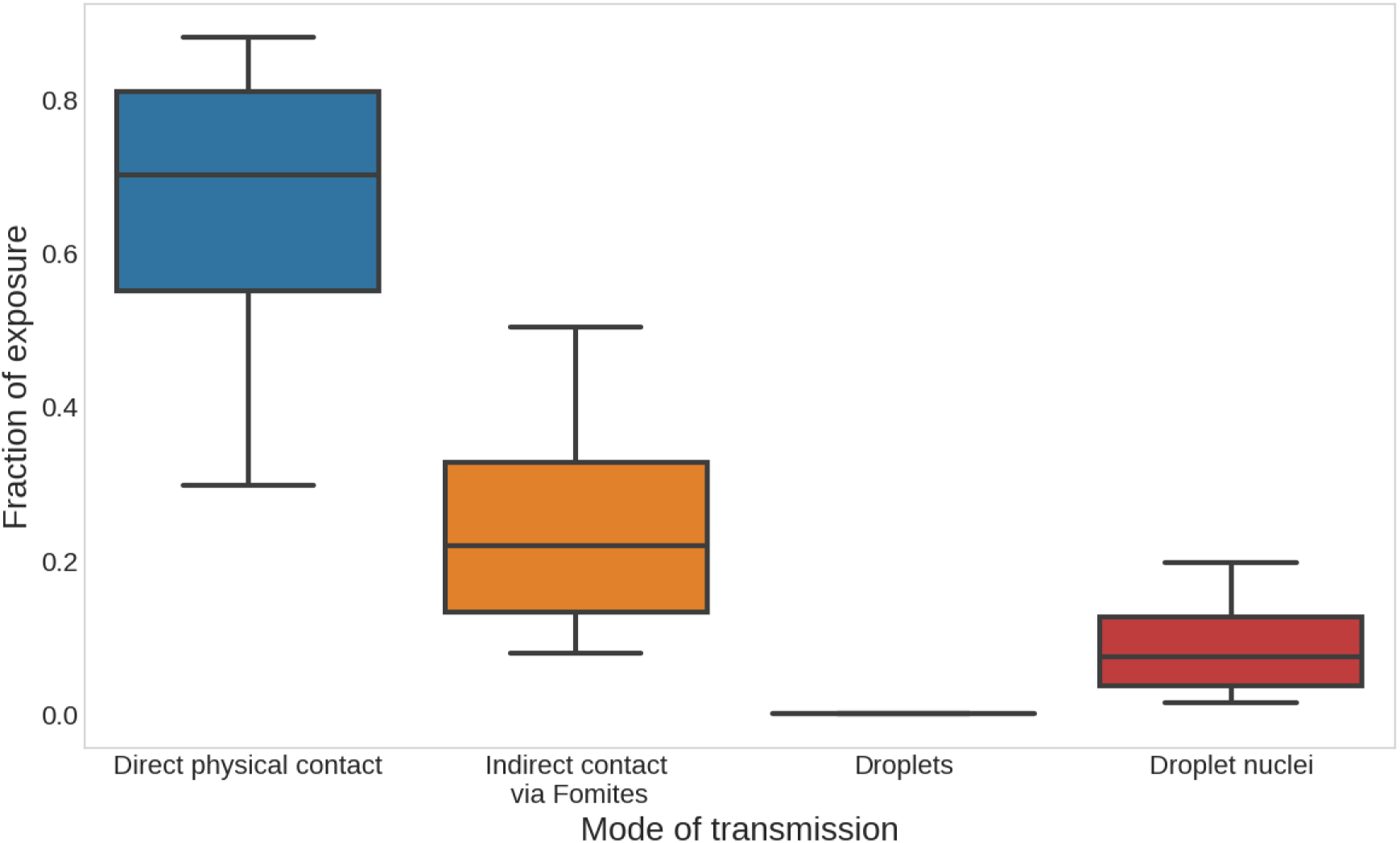
The Contribution of the different modes of transmission to overall exposure. Box represents the inter-quartile range (IQR). The whiskers represent the 10th and 90th percentile.

### Reducing the risk of infection

The fact that contact transmission is the main route of pre-symptomatic transmission, suggests that the hygienic and behavioral measures (HBMs) advised to the public should focus on reducing the contamination on the hands or somehow interrupt the hand-face transfer. We have examined five HBMs: Washing hands, cleaning fomites, avoiding physical contact (i.e., maintaining social distancing), wearing a mask and gloves. Naturally, conservative precautions measures would be an implementation of all these measures simultaneously. However, strict adherence to all these HBMs would be difficult to endure and to maintain over a long period of time. Therefore, we have tried to sort out several combinations of HBMs that should be readily implemented by the public, while significantly lowering the risk of infection. As the SAR is a proportion, it is appropriate to compare the HBMs in terms of odds ratio (*OR*), i.e., the odds that the secondary would be infected when a given combination of HBMs is taken, compared to the reference scenario in which no HBM is applied. Generally, any HBM that results in *OR* less than 1 decreases the risk of infection (i.e., provide smaller SAR than the reference)^34^. However, in practice the lower the *OR*, the more effective the HBM combination is at lowering the risk. The values brought here are in terms of *OR* alongside with 95% confidence interval (95% CI).

Washing hands is known to remove (and also destroy) virus particles from the hands and it is the simplest measure to implement. Our simulations show that washing hands once every hour rather than 3 times a day, as in the reference simulation (Table 1), results in *OR* of 0.72 (95% CI 0.67-0.8) (Figure 4A, column H). This result is consistent with intervention studies that have shown that increased hand washing decreased respiratory illness by 20%, albeit different viruses were studied^22^. This phenomenon seems counter intuitive, as we found that 90% of the viral dosage is transmitted through the hands and it was expected that washing it would remove the contamination. In order to understand the reason for the relatively limited effect of hand hygiene, we have examined the dynamics of the virus concentration on the hands of the secondary individual (Figure 4B). This concentration exhibits a periodic behaviour, with a period of approximately 30 to 40 minutes, that is governed by contact events on fomites and the face. Therefore, hand washing is expected to dramatically reduce the risk for infection if it occurs at at frequency higher than 40 min. Unfortunately, such frequent hand washing is unrealistic.

**Figure 4.**
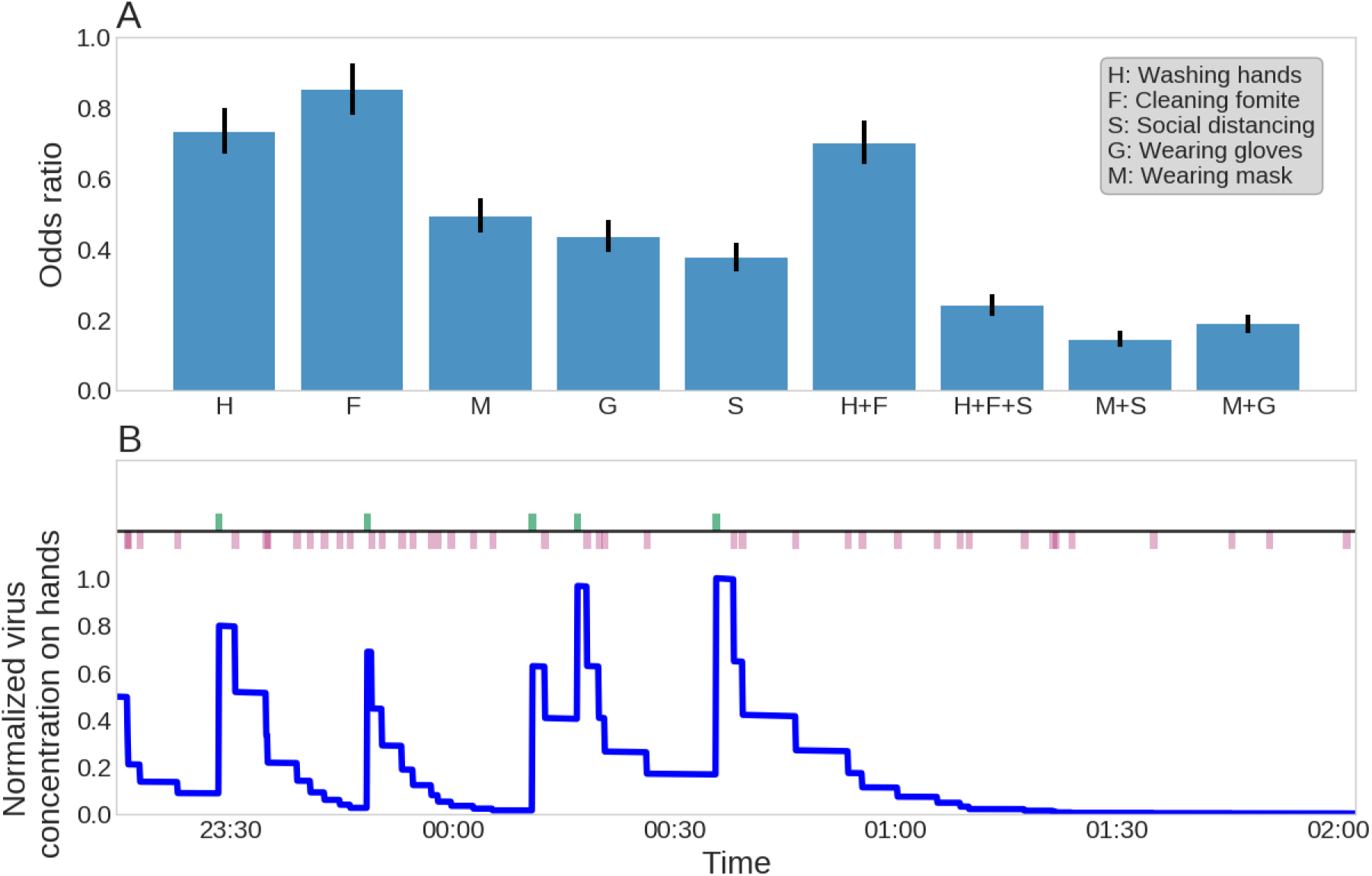
(A) The effect of hygiene and behavior on the risk of infection. Bars represent the confidence interval. (B) The normalized virus concentration over time. Green ticks represent fomite touching event. Red ticks represent face touching event.

Cleaning the fomites more frequently reduces the virus repositories that are available. Cleaning of the fomites 10 times a day rather than twice a day, as in the reference simulation, results in *OR* of 0.84 (95% CI0.77 −0.92), similar to washing hands more frequently (Figure 4A, column F). A combined strategy that consists of frequent hand washing and cleaning fomites does not decrease the risk considerably and results in *OR* of 0.70(95% CI 0.63 − 0.76).

Wearing a surgical mask or a respirator may reduce the hand-face transfer of virus particles^35^ as well as the inhalation exposure to viral particles. Although it is difficult to asses the reduction of the transfer coefficient from hand to facial membranes due to the use of a mask or a respirator, there are measurements regarding the protection provided against airborne transmission of bacteria and viruses. Available experimental results on N95 filtering face-piece respirators and surgical masks reported a protection factor of 2 − 10 for aerosols^36^. Hence, we have used a reduction of factor 2 in the hand-face transfer as well as in the exposure to airborne virus particles as a conservative estimate. With this value of the parameters, the simulated *OR* was 0.49 (95% CI 0.44 − 0.54) (Figure 4A, column M). Wearing gloves reduces the concentration of virus on the hands since the transfer efficiency from and to surfaces is halved with latex gloves^37^. As a result the use of gloves resulted *OR* of 0.43 (95% CI 0.39 − 0.48) (Figure 4A, column G).

Avoiding physical contact interrupts the main route of transfer between the two individuals and leads to an *OR* of 0.37 (95% CI 0.33 − 0.41) (Figure 4A, column S). Compared to the *OR*s resulted by taking a single HBM, this is the most effective step. A combined strategy that includes wearing masks and avoiding contact results in *OR* of 0.14 (95% CI 0.12-0.16, Figure 4A, column M+S), which is lower than wearing a mask and gloves (*OR* of 0.18, 95% CI0.16 − 0.21, Figure 4A, column M+G) or frequent hand washing, cleaning fomites and avoiding physical contact (*OR* of 0.23, 95% CI 0.21-0.27, Figure 4A, column H+F+S). The above shows that wearing masks and avoiding physical contact is the most effective HBM. However, wearing masks for long periods of time is difficult. Nevertheless, cleaning fomites, washing hands, and avoiding physical contact also provides considerable reduction in the *OR*. Hence, implementing these HBMs meticulously may save people the discomfort and limitation, that are associated with having to wear a mask constantly.

## Discussion

We have analyzed the possible routes of pre-symptomatic transmission in household scenarios. Using a validated model, we were able to identify the main mode of transmission as contact associated, mostly direct contact, but also contact mediated by fomites. The principal element in this transmission is hand-face transfer. Frequent hand washing and fomite cleaning coupled with the avoidance of physical contact result in a protection similar as wearing gloves and a mask. Although the present work does not account for highly populated indoor environments (such as work or commercial spaces), the relative importance of different processes is expected to remain very similar. However, in scenarios where people are in close proximity to each other (mass transit or during medical procedures), the contribution of the droplets to the transmission is expected to be larger. Hence, such scenarios require further investigation.

The relative contribution of the airborne transmission route is currently under debate. Several theoretical investigations concluded that the contribution of the airborne transmission route is significant^8, 9^. These studies assume that the viral load in the expelled droplets due to cough, sneeze and talking is 10^8^ − 10^9^ viruses/ml which is considerably higher than the reported median value^38^ of 7.5 · 10^5^ viruses/ml that was used in this work. In order to examine the effect of this assumption we used our model with the same maximal viral load of 10^8^ − 10^9^ viruses/ml. This resulted in a significant overestimation of the SAR. Specifically, we obtained a SAR of 70%, and to a SAR of 36% when only the airborne route was considered.

Our analysis, as with all modeling exercises, has several limitations and requires certain assumptions. The model does not account for contact patterns that prevail in households with young children and does not take into account the diurnal cycle of activity. The model parameters, such as the dose response curve, the viral shedding coefficients and transfer coefficients were chosen on the basis of knowledge of the SARS, other strains of coronavirus, or other bacteria^26, 39^. Although the model is stable with regard to variations in these parameters, more information on the key characteristic of the disease may reduce some uncertainties.

In conclusion, our findings can provide an important tool for decision makers while advising the public of the HBMs that are necessary to impede the progression of the epidemic. As it seems, recurrent outbreaks are expected to occur, as many countries will have to establish a fine balance between posing restrictions on society and allowing citizens to lead their life as normally as possible^40^. Under such a reality, the model presented in this study can be used to quantify the contribution of different HBM measures in order to devise guidelines that mitigate the risk of infection in scenarios of workplaces or schools, sports and cultural events, and mass transportation.

## Methods

In this section we delineate the mathematical details of the indoor transmission model that was developed in this study. The model simulates the transmission of the SARS-CoV-2 virus between two individuals that share the same indoor space, say, a room in a household or an office, using an agent based modeling approach. Specifically, it explicitly tracks the health condition of each individual, his actions, and the contamination in the indoors environment over time. The actions of the agents are randomized and therefore multiple realizations are required in order to obtain the appropriate ensemble statistics. At the beginning of the each realization, one individual, the primary, is infected but is pre-symptomatic. The other individual, the secondary, is susceptible.

The dynamics of the model are driven by the following processes:

1. Inoculation. Individual hands, which could be contaminated with viruses, touch the mouth and other facial membranes and exchange viruses with them. We denote 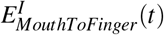 as the number of virus particles that pass from the facial membranes to the fingers of individual *I* (*I* is either primary or susceptible) after a single contact at time *t*,

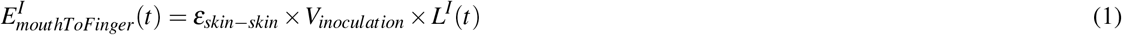

where *ε_skin_*_−_*_skin_* is the fraction of viruses that transfer in a skin to skin contact, *L*(*t*) is the current viral load of individual I at time *t* and *V_inoculation_* is the volume that carried from the facial membranes by the touch. The number of viruses that pass from the fingers to the facial membranes is denoted as *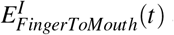* and equals to

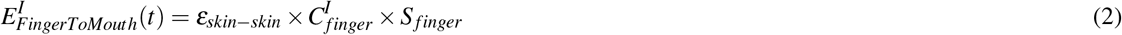

where 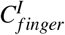 is the concentration of the finger and *S_finger_* is the effective surface area of the finger. Wearing gloves reduces the skin-skin transfer coefficient by a factor of 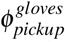 and wearing masks reduces the exchange of viruses between the facial membranes and the fingers by a factor of 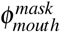
2. Coughing, sneezing, and talking. Individuals that cough, sneeze and talk emit particles within the range of 1*µm* to 2000*µm*,^22, 41, 42^. Infectious individuals emit particles that are loaded with viruses and thereby contaminate the nearby surfaces and the air. Particles greater than 100*µm*^3^ are referred to as large droplets. These droplets travel up to approximately 1.5m ^3^ before they settle on environmental surfaces. For simplicity, we assume that the contaminated area created by the large droplets from a single cough, sneeze, and talk is a semicircle with radius of 1.5m. The deposition of droplets directly on the mocusa of close contacts is a rare event in workplace or household settings^21^. Therefore, we have considered here only the contamination of environmental surfaces by the droplets after they have settled. Particles that are smaller than 100*µm* evaporate before they reach the ground and remain suspended in the air as droplet nuclei^3^. The droplet nuclei then disperse and might eventually infect a susceptible individual when he inhales them. Approximately 99% of the emitted particle volume following a sneeze, cough or talking is in droplets whose diameters are larger than 100*µm*^22, 41^. We assume that wearing masks completely blocks the emission of droplets and droplet nuclei.
3. Physical contact. Individuals also exchange viruses via physical contact with contaminated body parts. For simplicity, we consider the transfer from and to the fingers of the individuals, since the fingers (and specifically fingertips) are expected to be the most contaminated, as they are in contact with fomites and the facial mocusa. The number of viruses that pass from the primary to the secondary is the product of the skin to skin transfer fraction *ε_skin_*_−_*_skin_*, the concentration on the fingers 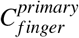 and the contact surface area *S_finger_*. Similarly, the transfer back to the primary is a product of *ε_skin_*_−_*_skin_* and *S_finger_* with the concentration on the fingers of the secondary 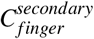. Therefore, the overall transfer between the primary and the secondary is

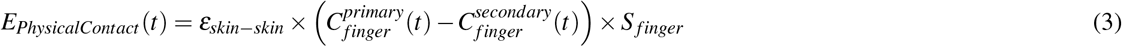 Wearing gloves reduces the skin-skin transfer coefficient by a factor of 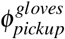
4. Fomite touching. Individuals touch fomite which results in virus exchanges between fomites and hands. Transfer efficiencies denoted by *ε_hand_*_−_ *_fomite_* (fomite to hand) and *ε_hand_*_−_ *_fomite_* (hand to fomite), were parameterized separately based on the results in Greene, et al.^43^, similar to the theoretical model of Kraay, et al.^28^. The number of viruses that transfers from the fomites to the hands is the product of the *ε_hand_*_−_ *_fomite_* with the the contact surface area (*S_finger_*) and the viable virus concentration on the fomite (*C_fomite_*). The number of viruses that transfers back to the fomite is the product of *ε_fomite_*_−_*_hand_*, *S_finger_* and the concentration on the finger. Therefore, the overall transfer from fomites to the hands following a single touch is given by,

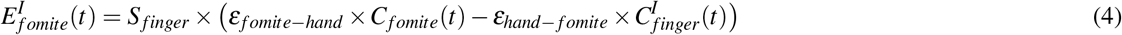

where *I* is the individual and is either primary or secondary. Wearing gloves reduces the fomite-hand and hand-fomite transfer coefficient by a factor of 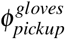, as above.
5. Touching surfaces. The large droplets that are emitted by coughs, sneezes, and speaks settle on environmental surfaces and contaminate them. When an individual touches an environmental surface his hand gets contaminated only if he touched a contaminated area. For simplicity, we assume that the probability to touch a given contaminated area is proportional to its size (*S_contaminatedArea_*) relative to the effective surface area of the room. The effective surface area is the sum of the room’s surface area (*S_room_*) and the furniture surface area (*S_furniture_*). The number of virus particles that are transferred to the hand with every touch is the product of the surface area of part of the hand that touches the surface *S_finger_*, the viable virus concentration on the surface *C_surface_* and the fraction that is transported to the mouth following that touch *ε_surface_*_−_*_hand_*. Accordingly,

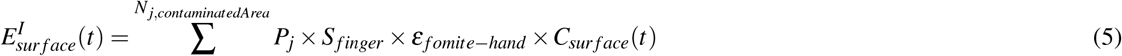 Where *P_j_* is the probability to touch a contaminated area on an environmental surface and given by the fraction of the contaminated area (*S_contaminatedArea_*) to the total surface area in the room, including the furniture (*S_room_* + *S_furniture_*). We neglect the transfer of virus particles back to environmental surfaces. Wearing gloves reduces the fomite-hand transfer coefficient by a factor of 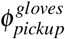, as above.
6. Hand washing. Washing hands removes the contamination from the hands of the individual with efficiency of *ε_washing_*. Hence, the number of viruses that is eliminated following a hand washing is

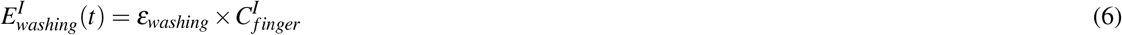
7. Probability to become infected. The probability to become infected is calculated from the virus dosage that was accumulated within each time interval. That is, we divide the simulation into time periods and calculate the overall exposure for each time period. The probability that a susceptible individual will become infected was inferred from the dose-response curve that was reported for SARS-CoV-1^26^,

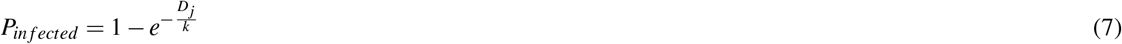

where *D_j_* is the total exposure of the individual at time period *j* (see Eqn 12 below). The length of each time period was determined from a Poisson process using a timescale of 7*h* that represents the life cycle of the virus in the body, inferred from the life cycle of the SARS-COV-1^44^.
8. Decay of the virus on surfaces. The viability of the virus on surfaces outside of a host body (such as fomites, environmental surfaces, and the hands) decays with time. The decay rate depends, generally, on the surface type and was inferred from measurements^45^.
9. Ventilation of contaminated air. The droplet nuclei that remain suspended are carried with the air currents in the room. For simplicity, we considered the entire house as a single, well mixed compartment. In households, the rooms are usually ventilated naturally (i.e., by the wind) and not by a heating, ventilation and air conditioning (HVAC) systems common in commercial and public buildings. The natural ventilation in households exchanges the air at a rate of 0.3*h*^−146, 47^, which leads to a decay of the air concentration of the droplet nuclei that carry the virus.

The above nine processes consist of both discrete random events (processes 1-7) and continuous events (processes 8 and 9). Hence, we used a hybrid continuous and stochastic-jump framework to model the dynamics of the transmission and infection processes.

The model consists of a system of coupled equations that describe the dynamics of the virus on the fingers of each individual, on fomites, in each contaminated area on an environmental area, its concentration in the air, and the overall exposure of the secondary individual from the airborne and contact routes. The concentration of the virus in each contaminated area on the environmental surface is tracked individually. A list of the parameters is provided in Table 2.

**Table 2.** The model parameters for the reference simulation

| Parameters | Parameter Description | Value | Unit | Reference |
| --- | --- | --- | --- | --- |
| SARS-CoV-2 specific parameters |  |  |  |  |
| $k$ | Dose-response coefficient | 410 | | 26 |
| $L_{max}$ | Maximal viral concentration in sputum | $7.5 \cdot 10^5$ | viruses/ml | 38 |
| $\epsilon_{cough}$ | Fraction of viral load emitted in cough | 0.001 | | 39 |
| $I_{mean}$ | Incubation period mean | 4.9 | day | 10, 24 |
| $I_{std}$ | Incubation period geometric std | 0.55 | | 10, 24 |
| $\tau_{exposure}$ | Time scale of the exposure | 6 | h | 44 |
| Individual parameters |  |  |  |  |
| B | Breathing rate | 10 | L/min |  |
| $\epsilon_{breath}$ | Fraction of breath exposure that lead to infection | 0.5 | | 50 |
| $S_{finger}$ | Surface area of a touch | 2 | $cm^2$ | 22 |
| $\tau_{PhysicalContact}$ | Rate of physical contacts in households | 3 | 1/d | 27 |
| $\tau_{inoculation}$ | Rate of face touching | 0.2 | 1/min | 22 |
| $\tau_{fomite}$ | Rate of fomite touching | 60 | 1/d | 28 |
| $\tau_{surface}$ | Rate of furniture touching | 1 | 1/min | 22 |
| $\tau_{hand-washing}$ | Rate of hand cleaning | 3 | 1/d | 28 |
| $\tau_{sneezing}$ | Rate of sneezing | 4 | 1/d | 51 |
| $\tau_{coughing}$ | Rate of coughing | 10 | 1/d | 52 |
| $\tau_{talking}$ | Rate of talking | 5 | 1/h | 53, 54 |
| $V_{cough, large}$ | Volume of cough droplets $> 100\mu m$ | 0.0598 | ml | 41 |
| $V_{cough, small}$ | Volume of cough droplets $< 100\mu m$ | $5.5 \cdot 10^{-4}$ | ml | 41 |
| $S_{contaminatedArea}$ | area of contaminated area on environmental surfaces | 3.5 | $m^2$ | 3, 22 |
| $V_{talk, large}$ | Volume of sneeze droplets $> 100\mu m$ | 0.0025 | ml | 41 |
| $V_{talk, small}$ | Volume of sneeze droplets $< 100\mu m$ | $3 \cdot 10^{-5}$ | ml | 41 |
| $V_{sneeze, large}$ | Volume of sneeze droplets $> 100\mu m$ | 4.35 | ml | 41 |
| $V_{sneeze, small}$ | Volume of sneeze droplets $< 100\mu m$ | 0.038 | ml | 41 |
| $V_{inoculation}$ | Volume of self inoculation | 0.01 | ml | 55 |
| $\epsilon_{fomite-hand}$ | fomite to hand transfer efficiency | 0.24 | | 37 |
| $\epsilon_{hand-fomite}$ | hand to fomite transfer efficiency | 0.05 | | 37 |
| $\epsilon_{skin-skin}$ | hand to hand transfer efficiency | 0.35 | | 22, 37, 56 |
| $\epsilon_{mouth}$ | Fraction of contact exposure that leads to infection | 0.5 | | 22 |
| $\epsilon_{washing}$ | efficiency of washing hands | 1 | | |
| $\alpha_{hand}$ | Virus decay rate on hands | 6 | 1/h | 45 |
| Room parameters |  |  |  |  |
| $S_{room}$ | Room surface area | 100 | $m^2$ | |
| $V_{room}$ | Room volume | 300 | $m^3$ | |
| $S_{furniture}$ | Furniture surface area | 80 | $m^2$ | 57 |
| $S_{fomit}$ | Fomite surface area | 13.3 | $cm^2$ | 58 |
| $\tau_{fomite-cleaning}$ | Rate of fomite cleaning | 2 | 1/d | 28 |
| $\alpha_{fomite}$ | Virus decay rate on fomite | 6 | 1/h | 45 |
| $\alpha_{furniture}$ | Virus decay rate on furniture | 6 | 1/h | 45 |
| $\alpha_{air}$ | Virus decay rate as aerosol | 1 | 1/h | 45 |
| $\beta$ | Air changes per hour | 0.3 | 1/h | 46, 47 |
| Hygienic and Behavioral measures |  |  |  |  |
| $\phi_{mouth}^{mask}$ | Masks hand-face transfer protection factor | 2 | | 36 |
| $\phi_{breath}^{mask}$ | Masks breathing protection factor | 2 | | 36 |
| $\phi_{pickup}^{gloves}$ | Gloves pickup protection factor | 2 | | 37 |

The dynamics of the concentration on the fingers of the individuals is determined by inoculation (process 1), physical contact (process 3), touching fomites (process 4), environmental surfaces (process 5), hand washing (process 6), and by the decay of the virus viability on the hands (process 8). Accordingly,

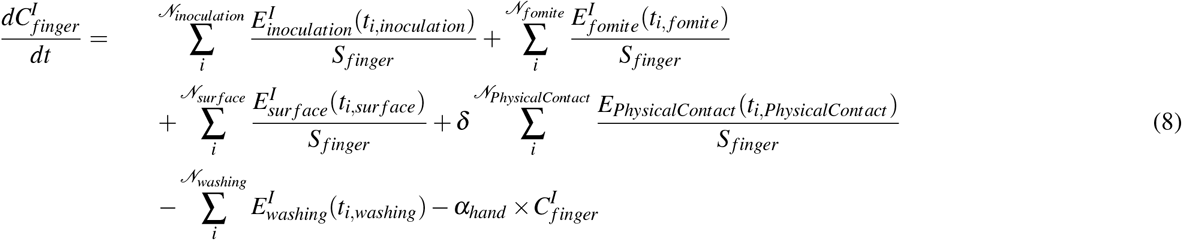

Where 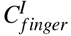 is the virus concentration on the fingers of individual I (I is either primary or secondary). The first 5 terms on the right hand side of Eqn. 8 describe the transfer of viruses as a result of the discrete events (processes 1,3,4,5, and 6) and the last term corresponds to the decay of the virus viability on the hands where *α_hand_* is the decay rate constant. The *δ* equals 1 for the primary individual and −1 for the secondary individual. The discrete event times (*t_i_*,*_X_* with *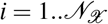* where X is inoculation, fomite, surface, washing and PhysicalContact) are determined from a Poisson distribution with rate constant *τ_X_* .

The dynamics of the average concentration of virus on the fomites in the room, *C_fomit_*, is determined by touching fomites (process 4) and the decay of the virus viability (process 8),

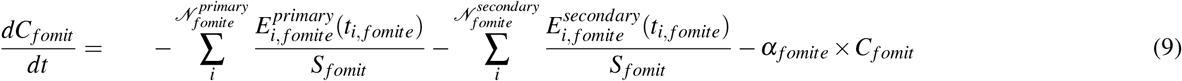

where *α_fomite_* is the decay rate in the fomite.

The dynamics of the concentration in the air is governed by the emission of droplet nuclei during the coughing, sneezing, and talking of the primary individual (process 2) and the ventilation process (process 9),

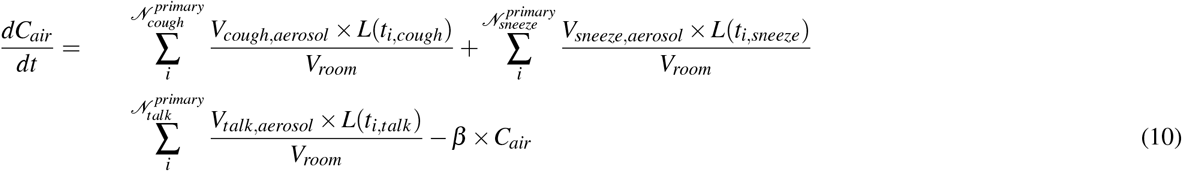

where *C_air_* is the virus concentration in the air and *L* is the viral load of the primary individual, 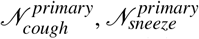 and 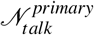 are the number of coughs, sneezes, and talks (respectively) that occur at times *t_i_*,*_cough_*, *t_i_*,*_sneeze_* and *t_i_*,*_talk_*, and determined from a Poisson distribution with time constants of *τ_cough_*, *τ_sneeze_*, and *τ_talk_*, respectively.

The dynamics of the concentration in the *k*-th contaminated area on an environmental surface that was created by a cough, sneeze or during a talk at time *T_k_* is determined by the equation

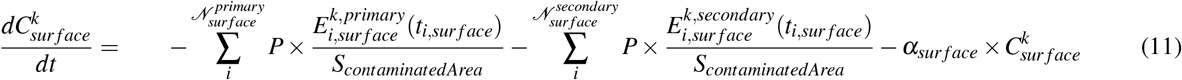

Where 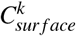 is the concentration on the *k*-th surface, *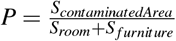* is the probability to touch a contaminated area and *α_surface_* is the decay rate in this surface.

The *k*-th contaminated area was created at time *T_k_* by a cough, sneeze or during speach. The initial concentration in the contaminated area is the product of the viral load of the primary individual at time *T_k_* with the volume of droplets that are larger than 100*µm* (*V_X_*,*_droplets_* where *X* is cough, sneeze or talk).

The exposure of the secondary is calculated within the exposure time interval and calculated as the sum of the exposure in the contact and droplet nuclei mode of transmission. Following Nicas and Best^22^, we assume that only a fraction *ε_mouth_* of the total number of virus particles that were deposited on the mouth reach oro-and nasopharyngel target sites. Similarly, only a fraction *ε_breath_* of the inhaled viruses deposit in the respiratory tracts. Therefore, the exposure is given by the equation:

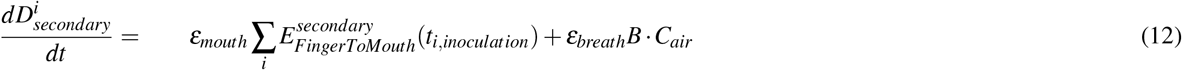

Where *D^i^* is the exposure obtained when solving the equation between the two exposure events that take place at times 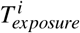 and 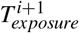 and *B* is the breathing rate.

### Numerical method

In order to solve the model’s equations 8 to 12 we chose to use the jump-adapted approximation proposed by Casella et al.^48^. For each realization, we determine the discrete event times (*t_i_*,*_X_* with 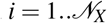 where X is inoculation, fomite, surface, washing, coughing, Physical Contact, sneezing and talking) are determined from a Poisson distribution with rate constant *τ_X_*. Then, the events are combined and sorted in an ascending order to obtain a set of discrete times. That is, 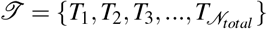, where each *T_i_* is assigned to a one *t_i_*,*_X_* event and 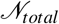 is the total number of discrete events.

The simulation is then solved implicitly between the times *T_i_* and *T_i_*_+1_ with time step of 6 seconds. At each *T_i_* we calculate the number of virus particles that is transferred for the corresponding event that takes place at this point in time.

The model was implemented in python version 3.6.5.

### Code availability

The computer code that was used in calculating the results of this paper is available in https://github.com/yehudarav/CoronaIndoorTransmission.git

### Parameter estimation

We have used results reported in the literature to determine the evidence based values for the parameters in the model. However, in some cases direct measurements were not available for several parameters. Therefore, their value was estimated, based on additional assumptions. In the following refer to these and we provide the details of these assumptions and their justifications.

Time scale of the exposure: Very little is known about the dynamics of the SARS-CoV-2 virus in the human body. In order to estimate the time scale of exposure, we used the results reported by Qinfen, et al.^44^ regarding the life cycle of the SARS-CoV-1 in host cells. They found that the virus assembly and maturation was first detected around 7 hours post infection. Thus, it is plausible to use this characteristic time as the exposure time interval.

Exposure to infection factor for contact and droplet nuclei mode of transmission: It is quite possible that not all the virus particles that are inhaled or reach the facial membranes cause infection. Therefore, we assumed, similar to Nicas and Best^22^, that the fraction of the exposure via the contact route that causes infection (*ε_mouth_*) is 0.5.

The fraction of the inhaled dose that causes infection was estimated to be roughly 0.5 of the deposition functions which provides the retention of particles in the lungs^49^ and the nasal cavity, depending on particle size^50^. These functions were applied to the particle size distribution reported by Chen, et al.^42^, after a correction that takes into account evaporation^3^.

Incubation time: We have used a weighted combination of the parameters of the incubation time distributions reported by Lauer, et al.^24^ and Bi, et al.^10^.

## Data Availability

Theoretical numerical model, all parameters are provided in the manuscript

## Acknowledgements

The authors would like to thank Dr. Shay Weiss and Dr. Yehudah Alexander for their fruitful comments. This research was not funded.

## Author contributions statement

All authors conceived the study and wrote the main manuscript. YA developed the model and did the simulation. EF contributed to numerical analysis. ZK contributed to parameter estimation. All authors contributed to the analysis and reviewed the manuscript.

## SUPPLEMENTARY INFORMATION

Theoretical investigation of pre-symptomatic SARS-CoV-2 person-to-person transmission in households

Yehuda Arav, Ziv Klausner, Eyal Fattal

### Sensitivity analysis

Some of the parameters’ values were obtained from studies that also reported the range of these values. Therefore, we have performed an extensive sensitivity analysis to check the robustness of the results. Specifically, we have examined the sensitivity of the model’s reconstruction of the serial interval distribution and the SAR for variations in the following parameters: dose response (Figure 5A), exposure time scale (Figure 5B), decay rate on surfaces (Figure 5C), surface area of the hand (Figure 5C), surface area touched (Figure 5D) and the median viral load (Figure 5E). As seen, the model’s results remains within the range of the values reported in the literature for the examined range of parameters.

**Figure 5.**
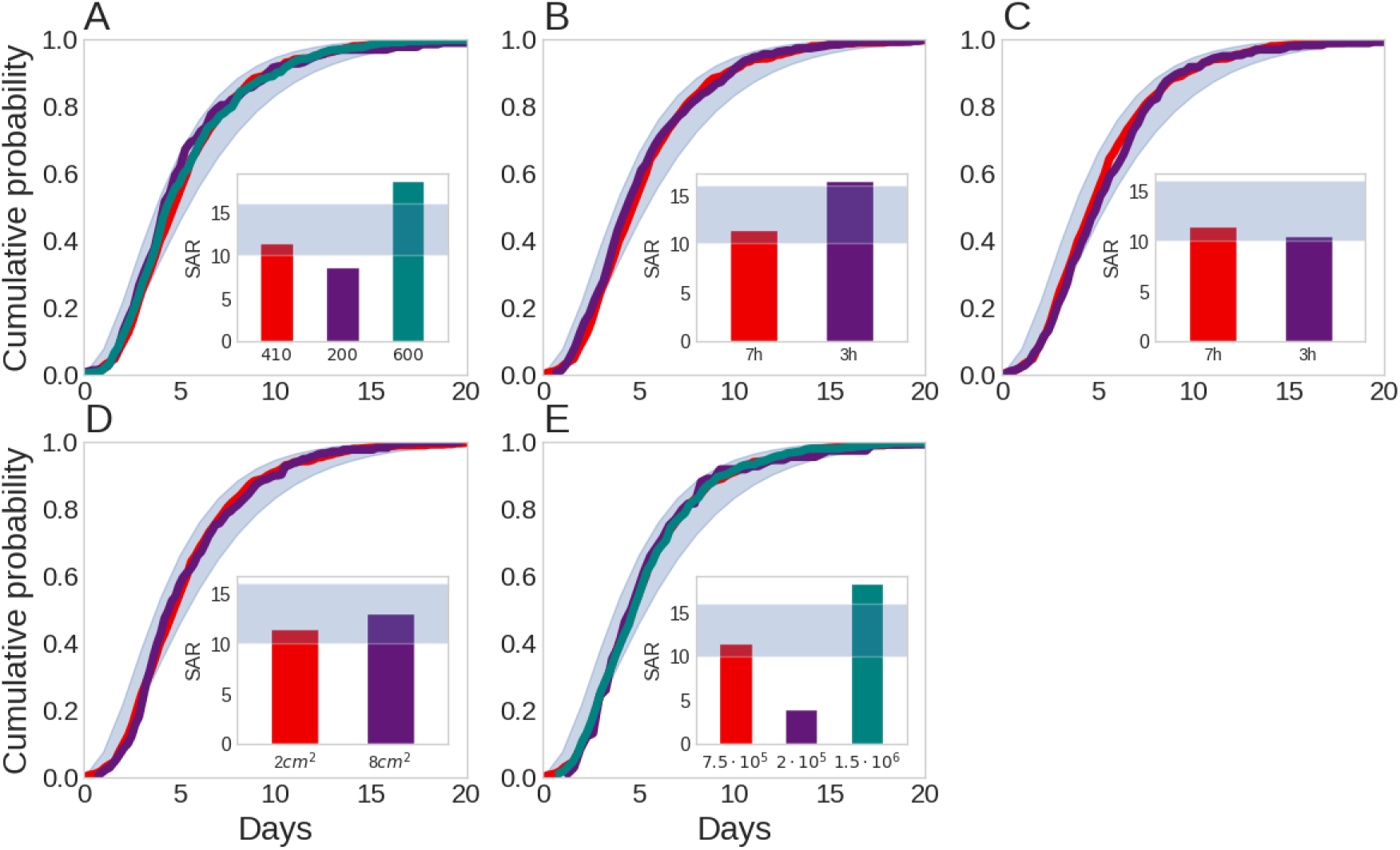
The prediction of the serial interval distribution and SAR (inset) for different parameter values. (A) dose response parameter (B) Exposure time scale (C) surface decay rate (D) surface area of a touch (E) median viral load. Red bar represents the reference simulation.

We note that since the contribution of transmission modes 3 and 4 (Indirect contact transmission via surface and droplet nuceli transmission, respectively) is very small, the model is not sensitive to the parameters that are associated with them (such as the area of the room, the decay rate in air, breathing rate and etc.).

## References

1. Gandhi, M., Yokoe, D. S. & Havlir, D. V. Asymptomatic Transmission, the Achilles’ Heel of Current Strategies to Control Covid-19. New Engl. J. Medicine 1–3, DOI: 10.1056/nejme2009758 (2020).

2. Kutter, J. S., Spronken, M. I., Fraaij, P. L., Fouchier, R. A. & Herfst, S. Transmission routes of respiratory viruses among humans, DOI: 10.1016/j.coviro.2018.01.001 (2018).

3. Xie, X. et al. How far droplets can move in indoor environments -revisiting the Wells ecaporation-falling curve. Indoor air 17, 211–225, DOI: 10.1111/j.1600-0668.2006.00469.x (2007).

4. Liu, J. et al. Community Transmission of Severe Acute Respiratory Syndrome Coronavirus 2, Shenzhen, China, 2020. Emerg. Infect. Dis. 26, 1320–1323, DOI: 10.3201/eid2606.200239 (2020).

5. Burke, R. M. et al. Active Monitoring of Persons Exposed to Patients with Confirmed COVID-19 -United States, January-February 2020. MMWR. Morb. mortality weekly report 69, 245–246, DOI: 10.15585/mmwr.mm6909e1 (2020).

6. Liu, J. et al. Community transmission of severe acute respiratory syndrome Coronavirus 2, Shenzhen, China, 2020. Tech. Rep. 6, World Health Organization (2020). DOI: 10.3201/eid2606.200239.

7. Li, Q. et al. Early Transmission Dynamics in Wuhan, China, of Novel Coronavirus-Infected Pneumonia. The New Engl. journal medicine 382, 1199–1207, DOI: 10.1056/NEJMoa2001316 (2020).

8. Vuorinen, V. et al. Modelling aerosol transport and virus exposure with numerical simulations in relation to SARS-CoV-2 transmission by inhalation indoors. Saf. Sci. 130, 104866, DOI: 10.1016/j.ssci.2020.104866 (2020). 2005.12612.

9. Buonanno, G., Stabile, L. & Morawska, L. Estimation of airborne viral emission: quanta emission rate of SARS-CoV-2 for infection risk assessment. medRxiv 2020.04.12.20062828, DOI: 10.1101/2020.04.12.20062828 (2020).

10. Bi, Q. et al. Epidemiology and Transmission of COVID-19 in Shenzhen China: Analysis of 391 cases and 1,286 of their close contacts. medRxiv 2020.03.03.20028423, DOI: 10.1101/2020.03.03.20028423 (2020).

11. Luo, L. et al. Modes of contact and risk of transmission in COVID-19 among close contacts. Digit. Econ. at Glob. Margins 2020.03.24.20042606, DOI: 10.7551/mitpress/10890.003.0026 (2020).

12. Park, S. Y. et al. Coronavirus Disease Outbreak in Call Center, South Korea. Emerg. Infect. Dis. 26, DOI: 10.3201/eid2608.201274 (2020).

13. Cheng, H.-Y. et al. High transmissibility of COVID-19 near symptom onset. medRxiv 2020.03.18.20034561, DOI: 10.1101/2020.03.18.20034561 (2020).

14. Santarpia, J. L. et al. Transmission Potential of SARS-CoV-2 in Viral Shedding Observed at the University of Nebraska Medical Center. medRxiv 2020.03.23.20039446, DOI: 10.1101/2020.03.23.20039446 (2020). 2020.03.23.20039446.

15. Chia, P. Y. et al. Detection of Air and Surface Contamination by Severe Acute Respiratory Syndrome Coronavirus 2 (SARS-CoV-2) in Hospital Rooms of Infected Patients. medRxiv 125, 2020.03.29.20046557, DOI: 10.1101/2020.03.29.20046557 (2020).

16. Dai, H. & Zhao, B. Association of infected probability of COVID-19 with ventilation rates in confined spaces: a Wells-Riley equation based investigation. medRxiv 2020.04.21.20072397, DOI: 10.1101/2020.04.21.20072397 (2020).

17. Faridi, S. et al. A field indoor air measurement of SARS-CoV-2 in the patient rooms of the largest hospital in Iran. Sci. Total. Environ. 725, 138401, DOI: 10.1016/j.scitotenv.2020.138401 (2020).

18. Ong, S. W. X. et al. Air, Surface Environmental, and Personal Protective Equipment Contamination by Severe Acute Respiratory Syndrome Coronavirus 2 (SARS-CoV-2) from a Symptomatic Patient, DOI: 10.1001/jama.2020.3227 (2020).

19. Döhla, M. et al. SARS-CoV-2 in environmental samples of quarantined households. medRxiv 49, 1–19, DOI: 10.1101/2020.05.28.20114041 (2020).

20. Nicas, M. & Sun, G. An integrated model of infection risk in a health-care environment. Risk Analysis 26, 1085–1096, DOI: 10.1111/j.1539-6924.2006.00802.x (2006).

21. Atkinson, M. P. & Wein, L. M. Quantifying the routes of transmission for pandemic influenza. Bull. Math. Biol. 70, 820–867, DOI: 10.1007/s11538-007-9281-2 (2008).

22. Nicas, M. & Best, D. A study quantifying the hand-to-face contact rate and its potential application to predicting respiratory tract infection. J. Occup. Environ. Hyg. 5, 347–352, DOI: 10.1080/15459620802003896 (2008).

23. Tankov, P. Financial Modelling with Jump Processes. Financial Model. with Jump. Process. DOI: 10.1201/9780203485217 (2003).

24. Lauer, S. A. et al. The Incubation Period of Coronavirus Disease 2019 (COVID-19) From Publicly Reported Confirmed Cases: Estimation and Application. Annals Intern. Medicine DOI: 10.7326/m20-0504 (2020).

25. He, X. et al. Temporal dynamics in viral shedding and transmissibility of COVID-19. medRxiv 2020.03.15.20036707, DOI: 10.1101/2020.03.15.20036707 (2020).

26. Watanabe, T., Bartrand, T. A., Weir, M. H., Omura, T. & Haas, C. N. Development of a dose-response model for SARS coronavirus. Risk Analysis 30, 1129–1138, DOI: 10.1111/j.1539-6924.2010.01427.x (2010).

27. Glass, R. J., Glass, L. M., Beyeler, W. E. & Min, H. J. Targeted social distancing design for pandemic influenza. Emerg. Infect. Dis. 12, 1671–1681, DOI: 10.3201/eid1211.060255 (2006).

28. Kraay, A. N. et al. Fomite-mediated transmission as a sufficient pathway: A comparative analysis across three viral pathogens. BMC Infect. Dis. 18, 540, DOI: 10.1186/s12879-018-3425-x (2018).

29. Fine, P. E. The Interval between Successive Cases of an Infectious Disease. Am. J. Epidemiol. 158, 1039–1047, DOI: 10.1093/aje/kwg251 (2003).

30. Wallinga, J. & Lipsitch, M. How generation intervals shape the relationship between growth rates and reproductive numbers. Proc. Royal Soc. B: Biol. Sci. 274, 599–604, DOI: 10.1098/rspb.2006.3754 (2007).

31. Du, Z. et al. The serial interval of COVID-19 from publicly reported confirmed cases. medRxiv 2020.02.19.20025452, DOI: 10.1101/2020.02.19.20025452 (2020).

32. Zhao, S. et al. Estimating the serial interval of the novel coronavirus disease (COVID-19): A statistical analysis using the public data in Hong Kong from January 16 to February 15, 2020. medRxiv 2020.02.21.20026559, DOI: 10.1101/2020.02.21.20026559 (2020).

33. Ganyani, T. et al. Estimating the generation interval for COVID-19 based on symptom onset data. medRxiv 2020.03.05.20031815, DOI: 10.1101/2020.03.05.20031815 (2020).

34. O’Connor, A. Interpretation of Odds and Risk Ratios. J Vet Intern Med 27, 600–603, DOI: 10.15537/smj.2019.12.24643 (2013).

35. Nicas, M. & Jones, R. M. Relative contributions of four exposure pathways to influenza infection risk. Risk Analysis 29, 1292–1303, DOI: 10.1111/j.1539-6924.2009.01253.x (2009).

36. Lee, S. A., Grinshpun, S. A. & Reponen, T. Respiratory performance offered by N95 respirators and surgical masks: Human subject evaluation with NaCl aerosol representing bacterial and viral particle size range. Annals Occup. Hyg. 52, 177–185, DOI: 10.1093/annhyg/men005 (2008).

37. Greene, C. et al. Fomite-fingerpad transfer efficiency (pick-up and deposit) of Acinetobacter baumannii -With and without a latex glove. Am. J. Infect. Control. 43, 928–934, DOI: 10.1016/j.ajic.2015.05.008 (2015).

38. Pan, Y., Zhang, D., Yang, P., Poon, L. L. M. & Wang, Q. Viral load of SARS-CoV-2 in clinical samples. The Lancet Infect. Dis. 20, 411–412, DOI: 10.1016/s1473-3099(20)30113-4 (2020).

39. Leung, N. H. L. et al. Respiratory virus shedding in exhaled breath and efficacy of face masks. Nat. Medicine 2020 1–5, DOI: 10.1038/s41591-020-0843-2 (2020).

40. Kissler, S. M., Tedijanto, C., Goldstein, E., Grad, Y. H. & Lipsitch, M. Projecting the transmission dynamics of SARS-CoV-2 through the postpandemic period. Science eabb5793, DOI: 10.1126/science.abb5793 (2020).

41. Duguid, J. P. The size and the duration of air-carriage of respiratory droplets and droplet-nuclei. J. Hyg. 44, 471–479, DOI: 10.1017/S0022172400019288 (1946).

42. Chen, S. C., Chio, C. P., Jou, L. J. & Liao, C. M. Viral kinetics and exhaled droplet size affect indoor transmission dynamics of influenza infection. Indoor Air 19, 401–413, DOI: 10.1111/j.1600-0668.2009.00603.x (2009).

43. Greene, C. et al. Fomite-fingerpad transfer efficiency (pick-up and deposit) of Acinetobacter baumannii -With and without a latex glove. Am. J. Infect. Control. 43, 928–934, DOI: 10.1016/j.ajic.2015.05.008 (2015).

44. Qinfen, Z. et al. The life cycle of SARS coronavirus in Vero E6 cells. J. Med. Virol. 73, 332–337, DOI: 10.1002/jmv.20095 (2004).

45. van Doremalen, N. et al. Aerosol and Surface Stability of SARS-CoV-2 as Compared with SARS-CoV-1. New Engl. J. Medicine DOI: 10.1056/nejmc2004973 (2020).

46. Hou, J. et al. Air Change Rates in Residential Buildings in Tianjin, China. In Procedia Engineering, vol. 205, 2254–2258, DOI: 10.1016/j.proeng.2017.10.069 (Elsevier Ltd, 2017).

47. Yamamoto, N., Shendell, D. G., Winer, A. M. & Zhang, J. Residential air exchange rates in three major US metropolitan areas: Results from the Relationship among Indoor, Outdoor, and Personal Air Study 1999-2001. Indoor Air 20, 85–90, DOI: 10.1111/j.1600-0668.2009.00622.x (2010).

48. Casella, B. & Roberts, G. O. Exact Simulation of Jump-Diffusion Processes with Monte Carlo Applications. Methodol. Comput. Appl. Probab. 13, 449–473, DOI: 10.1007/s11009-009-9163-1 (2011).

49. Carvalho, T. C., Peters, J. I. & Williams, R. O. Influence of particle size on regional lung deposition -What evidence is there? Int. J. Pharm. 406, 1–10, DOI: 10.1016/j.ijpharm.2010.12.040 (2011).

50. Breysse, P. N. & Swift, D. L. Inhalability of large particles into the human nasal passage: In vivo studies instill air. Aerosol Sci. Technol. 13, 459–464, DOI: 10.1080/02786829008959460 (1990).

51. Hansen, B. & Mygind, N. How often do normal persons sneeze and blow the nose?*. Rhinology 10–12 (2002).

52. Hsu, J. Y. et al. Coughing frequency in patients with persistent cough: Assessment using a 24 hour ambulatory recorder. Eur. Respir. J. 7, 1246–1253, DOI: 10.1183/09031936.94.07071246 (1994).

53. Jiahong, Y., Liberman, M. & Cieri, C. Towards an integrated understanding of speaking rate in conversation. Proc. Annu. Conf. Int. Speech Commun. Assoc. INTERSPEECH 2, 541–544 (2006).

54. Mehl, M. R., Vazire, S., Ramírez-Esparza, N., Slatcher, R. B. & Pennebaker, J. W. Are women really more talkative than men? Science 317, 82, DOI: 10.1126/science.1139940 (2007).

55. U.S. Environmental Protection Agency. USEPA Exposure Factors Handbook Chapter 7: Dermal Exposure Studies. Expo. Factors Handb. (2011).

56. Rusin, P., Maxwell, S., Microbiology, C. G. J. o. A. & undefined 2002. Comparative surface-to-hand and fingertip-to-mouth transfer efficiency of gram-positive bacteria, gram-negative bacteria, and phage. Wiley Online Libr. 585–592 (2002).

57. Manuja, A. et al. Total surface area in indoor environments. Environ. Sci. Process. Impacts 21, 1384–1392, DOI: 10.1039/c9em00157c (2019).

58. Beamer, P. I. et al. Modeling of human viruses on hands and risk of infection in an office workplace using micro-activity data. J. Occup. Environ. Hyg. 12, 266–275, DOI: 10.1080/15459624.2014.974808 (2015).

